# Quantifying the potential value of antigen-detection rapid diagnostic tests for COVID-19: a modelling analysis

**DOI:** 10.1101/2020.11.20.20235317

**Authors:** Saskia Ricks, Emily A. Kendall, David W. Dowdy, Jilian A. Sacks, Samuel G. Schumacher, Nimalan Arinaminpathy

**Affiliations:** MRC Centre for Global Infectious Disease Analysis, Imperial College London, UK; Division of Infectious Diseases, Johns Hopkins University School of Medicine, Baltimore, MD, USA; Department of Epidemiology, The Johns Hopkins Bloomberg School of Public Health, Baltimore, MD, USA; Foundation for Innovative New Diagnostics, Geneva, Switzerland

## Abstract

**Background:** Testing plays a critical role in treatment and prevention responses to the COVID-19 pandemic. Compared to nucleic acid tests (NATs), antigen-detection rapid diagnostic tests (Ag-RDTs) can be more accessible, but typically have lower sensitivity and specificity. By quantifying these trade-offs, we aimed to inform decisions about when an Ag-RDT would offer greater public health value than reliance on NAT.

**Methods:** Following an expert consultation, we selected two use cases for analysis: rapid identification of people with COVID-19 amongst patients admitted with respiratory symptoms in a ‘hospital’ setting; and early identification and isolation of people with mildly symptomatic COVID-19 in a ‘community’ setting. Using decision analysis, we evaluated the cost and impact (deaths averted and infectious days isolated) of an Ag-RDT-led strategy, compared to a strategy based on NAT and clinical judgment. We performed a multivariate sensitivity analysis to identify key parameters.

**Results:** In a hospital setting, an Ag-RDT-led strategy would avert more deaths than a NAT-based strategy, and at lower cost per death averted, when the sensitivity of clinical judgement is less than 85%, and when NAT results are available in time to inform clinical decision-making for less than 90% of patients. The use of an Ag-RDT is robustly supported in community settings, where it would avert more transmission at lower cost than relying on NAT alone, under a wide range of assumptions.

**Conclusions:** Despite their imperfect sensitivity and specificity, Ag-RDTs have the potential to be simultaneously more impactful, and cost-effective, than current approaches to COVID-19 diagnostic testing.

## Introduction

Virological testing is a critical part of the global response to SARS-CoV-2(1–3). Early diagnosis allows infectious cases to be isolated in a timely manner, thus minimising opportunities for transmission. Amongst those at risk of severe outcomes of the disease, early diagnosis and initiation of appropriate therapy can substantially improve outcomes and avert mortality (4–7). Nucleic acid tests (NATs) have been widely implemented in well-resourced settings since the outset of the pandemic and have the benefit of high sensitivity and specificity for current or recent infection. However, these tests are challenging to implement at scale, particularly in resource-poor settings: they are costly and require good specimen transport systems, laboratory infrastructure, and highly trained technicians. Delays of a week or more in obtaining results after collecting specimens are therefore common (8–10), and in such cases a NAT result adds little value to decisions around isolation or clinical management.

The emergence of antigen-detection rapid diagnostic tests (Ag-RDTs) may help to address some of these challenges. The WHO have recently published Target Product Profiles for such tests (11), which detect SARS-CoV-2 proteins (antigens) to diagnose active infection. Ag-RDTs can be conducted relatively easily, at low cost, and within minutes, at the point of care without need for a laboratory. However, they have lower sensitivity and specificity and may miss SARS-CoV-2 in specimens with lower quantities of virus. For example, available Ag-RDTs were estimated to have less than 80% sensitivity for COVID-19, compared with >90% for NAT (12). We therefore sought to quantify these trade-offs between Ag-RDT-based testing and NAT-based testing in the context of resource-limited settings.

## Methods

### Overview

Our primary objective was to identify scenarios in which an Ag-RDT might offer greater individual and public health value at lower cost than reliance on NAT. To accomplish this, we first defined key use cases and plausible ranges for parameter values in consultation with a group of experts deeply involved in their country’s response to COVID-19. We then constructed decision trees that included both costs and relevant outcomes (deaths and infectious person-days averted). Finally, we simulated overall costs and outcomes under a wide array of parameter values and compared testing strategies using Ag-RDTs, those using NAT where available. We also constructed a user-friendly online tool (https://covid-ag-rdt.shinyapps.io/model/) that enables public health practitioners to examine model outputs for input parameter values relevant to their own settings.

#### Model scenarios and structure

We denote an ‘Ag-RDT-led strategy’ as any testing strategy in which an Ag-RDT is the first diagnostic test performed (with the potential for follow-up NAT confirmation). As an illustrative example, we focused on an Ag-RDT with sensitivity and specificity of 80% and 98% respectively and costing 5 US$ per test, consistent with recent WHO interim guidance and US FDA emergency use authorized antigen-detection tests (13,14). We compared the impact of using an Ag-RDT-led strategy to that of a ‘NAT-based strategy’ in which NAT was the only virological test performed, with reliance on clinical judgment where sufficiently rapid NAT results were not available (see Fig. 1 for a summary of the diagnostic strategies modelled). To inform relevant use case scenarios, we consulted experts from India, South Africa, Nigeria and Brazil to elicit expert opinion on the ways in which Ag-RDTs could offer value in their own country settings (see supporting information for further details). Based on this input, we selected two use case scenarios, as listed in Table 1: (i) A ‘hospital setting’, where the test is used to support infection control and treatment decisions amongst patients being admitted to hospital with respiratory symptoms, and (ii) a ‘community’ setting, where the test is used in in decentralized community clinics to identify cases of COVID-19 who should self-isolate. Although Ag-RDTs could also be considered for use in identifying asymptomatic infections, both of these focal scenarios involved testing of only symptomatic individuals.

**Table 1.** Two use cases included in the present analysis of the relative value of antigen-detection rapid diagnostic tests (Ag-RDTs) compared to nucleic acid tests (NATs).

| Use case scenario | Description | Assumed prevalence | Purpose of testing |
| --- | --- | --- | --- |
| Hospital setting | Testing amongst all patients being hospitalised with respiratory symptoms | 25% | To identify patients with COVID-19 who should be housed in isolation wards (reducing infection risk in hospital)<br>Amongst admissions with severe symptoms, to identify those with COVID-19 who might benefit from anti-inflammatory treatment (to reduce deaths) |
| Community setting | Decentralised, community-level facility available to all individuals with symptoms who want to be tested for COVID-19 | 5% | To identify COVID-19 amongst people with mild COVID-consistent symptoms. Positive test results would trigger isolation and contact tracing, to minimise opportunities for transmission. |

**Figure 1.**
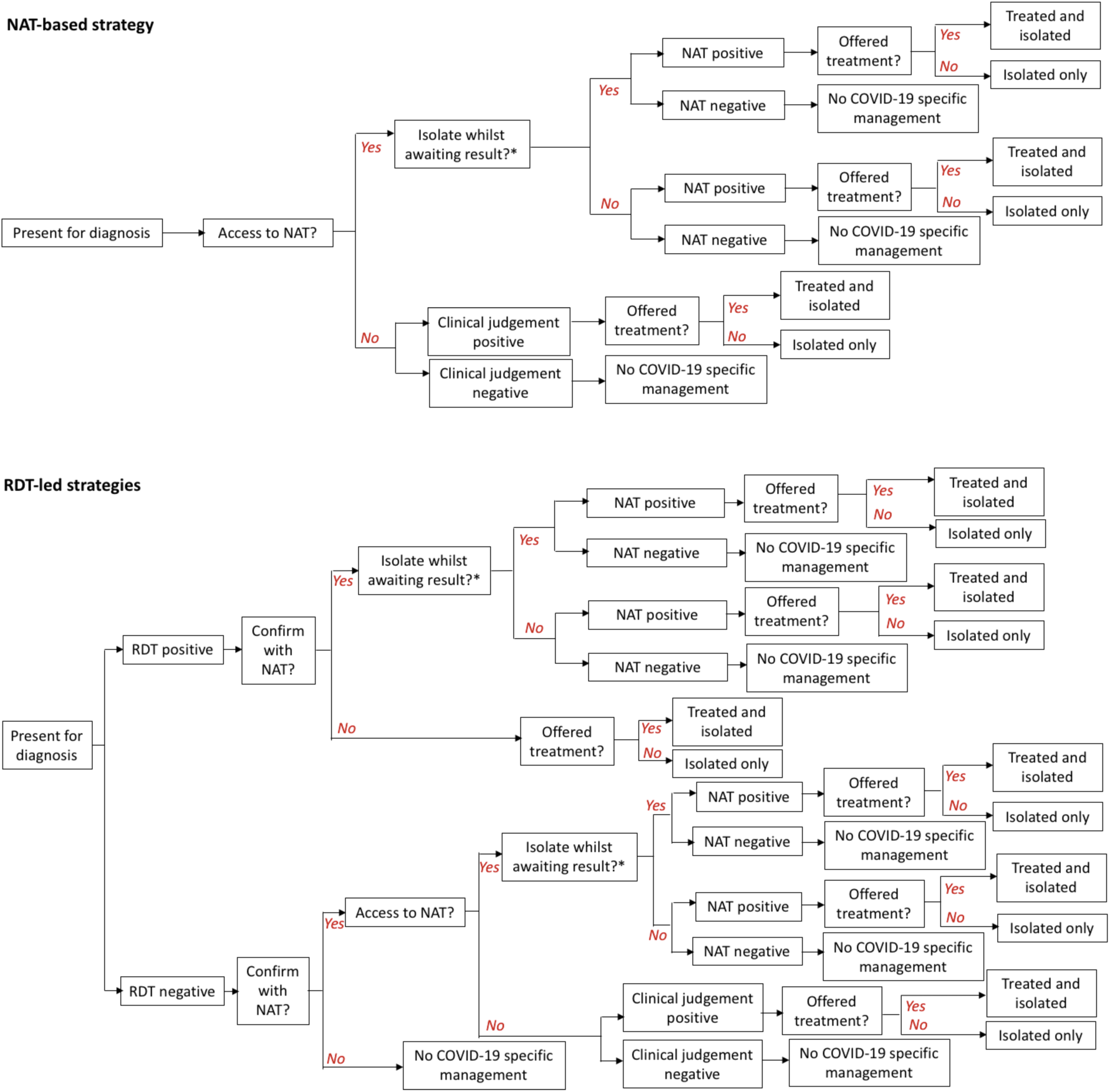
Schematic illustration of the decision tree approach for modelling the cost and impact of different testing strategies. As described in the main text, our analysis focuses on the direct benefit to patients being tested in different settings. *In addition to isolating whilst awaiting a NAT result, patients with severe SARS-CoV-2 are also started on treatment. Costs and deaths/infectious days averted are accumulated along each branch of the diagram as appropriate (for example, counting the costs of interim treatment and isolation along any branch labelled ‘Yes’ following ‘Isolate whilst awaiting treatment?’).

For both use cases we constructed decision trees (Figure 1) that represent the diagnostic use of the Ag-RDT, actions taken in response to the test results (or lack of results), and resulting outcomes. For simplicity and transparency, this model does not incorporate transmission dynamics but approximates epidemiological benefits based on the incremental change in the number of days that infectious individuals spend out of isolation; the magnitude of downstream impact would depend on factors, such as the rate of epidemic growth and the contact patterns of symptomatic versus pre- or asymptomatic cases, that are not specified in our model. Our focus is therefore on the direct benefits that would accrue to patients receiving the test and, by extension, their immediate contacts (see right-hand column of Table 1).

Model parameters, listed in Table 2, represent the contextual factors to be examined (including plausible ranges for each), with the aim of identifying those factors that are most influential for the value of an Ag-RDT-led testing strategy relative to NAT-based testing. Our expert consultation highlighted that no standard guidance for whether or how Ag-RDTs should be used in conjunction with NAT existed at the time (e.g. whether NAT should be used to confirm an Ag-RDT negative result). Thus, we also defined and modelled three different options for the adjunctive use of NAT in an Ag-RDT-led algorithm: (i) no confirmation of Ag-RDT results; (ii) NAT confirmation of Ag-RDT negative results; or (iii) NAT confirmation of Ag-RDT positive results).

**Table 2.** Contextual parameters and their uncertainty ranges. For each parameter, we assumed a uniform distribution spanning the range shown. Footnotes: *We exclude any parameter draws involving NAT confirmation of *both* Ag-RDT negative and Ag-RDT positive results. **For patients with COVID-19 initiated on interim treatment while awaiting NAT confirmation of an Ag-RDT result, but who subsequently receive an incorrect negative NAT result, we assume that they do not receive any benefits of interim treatment.

| Parameter | Value |  | References |
| --- | --- | --- | --- |
|  | Hospital setting | Community setting |  |
| **Epidemiology** |  |  |  |
| Prevalence of current or recent SARS-CoV-2 infection (%) | 25 | 5 | Assumption |
| Proportion amongst those tested who are in acute phase | 0.5– 1.00 | 0.5– 1.00 | Assumption |
| Of those in acute phase, number of infectious days remaining (days) | 5 – 15 | 5 – 15 | (27) |
| Proportion who would die without treatment | 0.05 – 0.10 | N/A | (27,28) |
| Amongst those who would die without treatment, reduction in mortality by treatment (1 indicates full reduction) | 0.40 – 0.60 | N/A | (4,7,29–31) |
| **NAT performance** |  |  |  |
| NAT sensitivity (for current or recent COVID-19) | 0.95 – 0.99 | 0.95 – 0.99 | (15–17) |
| NAT specificity | 0.99 – 1 | 0.99 – 1 | (15–17) |
| NAT availability (proportion able to access NAT test) | 0.1 – 1 | 0.1 – 1 |  |
| Cost per NAT test (\$) | 20 – 70 | 20 – 70 | |
| NAT turnaround time (days) | 1 – 10 | 5 – 15 | (10), Expert consultation |
| Confirm Ag-RDT negative results with NAT | Y/N | Y/N |  |
| Confirm Ag-RDT positive results with NAT * | Y/N | Y/N |  |
| Isolate and initiate treatment (if indicated) whilst awaiting NAT result | Y ** | N |  |
| **Ag-RDT performance (assumed fixed)** |  |  |  |
| Ag-RDT sensitivity for current infection (% , assumed only amongst acute cases) | 0.80 | 0.80 | (13,14) |
| Ag-RDT specificity (%) | 0.98 | 0.98 | (13,14) |
| Cost per Ag-RDT test (\$) | 5 | 5 | |
| **Clinical judgement and management** |  |  |  |
| Sensitivity of clinical judgment in absence of NAT | 0.45 – 0.99 | 0.45 – 0.99 | (32–35) |
| Specificity of clinical judgment in absence of NAT | 0.20 – 0.50 | 0.20 – 0.50 | (32–35) |
| Amongst those who would survive without treatment (including COVID -ve and COVID +ve), | 0 – 0.1 | N/A |  |
| proportion erroneously identified as needing treatment |  |  |  |
| Amongst COVID +ve who would die without treatment, proportion correctly identified as such | 0.8 – 1 | N/A |  |
| Duration of isolation (days) | 10 | 10 | (27) |
| Duration of treatment (days) | 10 | N/A | (7) |
| Cost of isolation per day (\$) | 10 – 100 | 10 – 100 | |
| Cost of treatment per day (\$) | 50 – 500 | N/A | (36,37) |

It is also necessary to specify scenarios for interim management of patients while awaiting a NAT result (whether in the NAT-based strategy or while awaiting NAT confirmation in an Ag-RDT-led strategy). In the hospital setting, we assumed that: (i) all patients are isolated while awaiting a NAT result, and (ii) those thought to need treatment are initiated on treatment during this time: in the supporting information we also present sensitivity analysis, to the alternative scenario where there is no isolation, nor treatment pending NAT results. By contrast, in the community setting we assumed that individuals are *not* isolated while awaiting any NAT result, as this policy was considered infeasible due to the comparatively low prevalence of COVID-19 in this population, and the unnecessary expense and disruption this would entail to most tested individuals and their families.

Although NAT specificity is near 100% for current or recent infection (15), not all NAT-positive cases are necessarily infectious, given the potential to detect unviable viral genetic material after the infection has resolved (15–17) and for severe symptoms to develop near the end of the infectious period (18). By contrast, Ag-RDTs may detect only acute, but not recently cleared, infection (19,20). These distinctions have significance for the intended purpose of the test: where the purpose is to guide clinical decisions for treatment, knowing the aetiology of severe symptoms is important, regardless of viral antigenic load. On the other hand, where the purpose is early identification of infectious cases, detecting recently-cleared infection can detract from the utility of a test. We captured these elements of both NATs and Ag-RDTs by distinguishing ‘acute’ from ‘recent’ infection, and assuming that: (i) only acute infection is infectious; (ii) NAT is able to detect both acute and recent infection with equal sensitivity, and (iii) an Ag-RDT is able to detect only acute infection (19). As discussed below, although these are useful simplifications for the purpose of the current analysis, these categorisations conceal potentially important complexities relating to temporal and between-individual variation in viral load, infectivity and detectability by a given test. In the present analysis we incorporated a parameter for the proportion of those with COVID-19, amongst the population being tested, that are still in the acute phase at the point of testing, allowing this parameter to occupy a wide range of values between 50% and 100%, acknowledging the existing uncertainty (See Table 2).

#### Quantifying relative value

To estimate the impact of a test on deaths in a hospital setting, we assumed that a certain proportion of admitted patients would die without treatment. We assumed that early treatment of these individuals would reduce mortality by 5 – 50% (Table 2), consistent with recent study results for corticosteroid treatment of COVID-19 (7). We assumed further that individuals with severe disease would be eligible for treatment while awaiting a test result, as well as once confirmed as having COVID-19. We thus denoted ‘deaths averted’ as the reduction in deaths that would be achieved by a given testing strategy, relative to no intervention. Similarly, as a simple proxy for the impact of a test on transmission, we first assumed a uniform distribution for the number of infectious days remaining per patient amongst patients presenting with acute infection (Table 2). We then recorded the number of patient-days of acute infection that were not spent in isolation, whether because of missed diagnosis or (in the case of NAT) delayed diagnosis without isolation, while awaiting a test result. We denoted ‘infectious person-days averted’ as the reduction that would be achieved by a given testing strategy, relative to no intervention.

Using the model illustrated in Figure 1, we estimated the impact (deaths or infectious days averted) and cost of each testing strategy. We stratified Ag-RDT-led strategies by the adjunctive role of NAT in confirmation of a test result (i.e. whether to confirm Ag-RDT-negatives, Ag-RDT-positives, or not at all). For NAT-based strategies, we assumed that only a proportion of eligible individuals receive a NAT result (assuming a broad range of 10-100%), with the remainder managed through clinical judgment alone. For each use case, we sampled all parameters from the uncertainty ranges in Table 2 using Latin Hypercube Sampling. For each sampled set of parameters, we calculated both incremental costs and the incremental primary outcome (deaths averted or infectious person-days that were isolated) under an Ag-RDT-led strategy or a NAT-based strategy, relative to no intervention (that is, a scenario of no testing, nor clinical management of COVID-19). To quantify uncertainty, we calculated uncertainty intervals (UIs) as 2.5^th^ and 97.5^th^ percentiles over 10,000 samples, and reported median values as point estimates.

To compare testing strategies, we first estimated the cost-per-death averted (in the hospital setting), and the cost-per-infectious person day isolated (in both hospital and community settings) under NAT-based and Ag-RDT-led strategies. However, we did not aim to determine whether or not an Ag-RDT would be cost-effective, given the uncertainties surrounding appropriate willingness-to-pay thresholds for emergency outbreak response (21). Instead, we compared the two strategies (Ag-RDT vs NAT) using a simple approach of plotting their relative impact against their relative cost for each sampled set of parameters (see Figure S1 for a schematic illustration of the approach). It is important to note that this approach is distinct from a conventional cost-effectiveness plane, as the axes are shown on a relative, rather than a nominal, scale. In the example of deaths, we denoted *A*_*RDT*_ as the deaths averted by Ag-RDT-led testing, relative to no intervention, and likewise for *A*_*NAT*_. Similarly, we calculated the incremental cost *C*_*RDT*_ of an Ag-RDT-led strategy relative to no intervention, and likewise for *C*_*NAT*_. We then plotted the relative impact (*A*_*RDT*_*/A*_*NAT*_) against the relative incremental cost (*C*_*RDT*_*/C*_*NAT*_).

We defined an Ag-RDT as being ‘favourable’ relative to NAT, wherever its use results simultaneously in more deaths averted than NAT (i.e. *A*_*RDT*_> *A*_*NAT*_), and a lower incurred cost per death averted than NAT (i.e. *C*_*RDT*_*/A*_*RDT*_< *C*_*NAT*_*/A*_*NAT*_). We defined an Ag-RDT as being ‘non-favourable’ otherwise. We performed corresponding calculations for the outcome of infectious person-days successfully isolated. Our focus in the following analysis is on identifying which circumstances would lead to an Ag-RDT being ‘favourable’ relative to NAT.

Where simulation outputs were equivocal on the favourability of Ag-RDTs, i.e. straddling favourable and non-favourable regions, we evaluated the correlation between each parameter and relevant model outputs using partial rank correlation coefficients, to identify those parameters that were most influential on the probability of a simulation falling in a favourable region. In particular, where simulation outputs straddled the vertical dashed line shown in Fig.S1, we evaluated correlations against the relative impact of Ag-RDT-led vs NAT-based testing strategies. Where simulations straddled the diagonal line in the upper-right quadrant, we evaluated correlations against the relative cost-per-unit impact (i.e. per death averted or per infectious person-day isolated). Overall, in this way we sought to identify the contextual conditions under which an Ag-RDT-led strategy would, and would not, be favoured over NAT.

## Results

Table S2 shows estimates of incremental cost-effectiveness ratios (relative to a scenario of no intervention), illustrating, for example, that – relative to no intervention – a NAT based algorithm in a hospital setting would cost $119,830 (95% uncertainty intervals (UI) 32,080 – 491,070) per death averted within the patient population, while an Ag-RDT-led algorithm, involving NAT confirmation of Ag-RDT-negatives, would cost $113,130 (29,470 – 451,600). Likewise, in a community setting, a NAT based algorithm would cost $732 (179 – 2,541) per infectious person-day isolated, while an Ag-RDT-only algorithm (without NAT confirmation) would cost only $88 (26 – 227) per infectious person-day isolated.

### Hospital Setting

Figure 2 shows plots of relative incremental cost against relative impact in terms of deaths averted in a hospital setting, with an assumed 25% prevalence of acute or recent COVID-19 amongst those being tested. For deaths averted, when an Ag-RDT is used in conjunction with NAT to confirm Ag-RDT-negative results (blue points), such a strategy had greater impact, and at lower cost per death averted, than a NAT-based strategy (“favourable region”) in 89% of all simulations. By contrast, Ag-RDT-led strategies that either involve no NAT confirmation, or only confirmation of RDT-positive cases (respectively orange and yellow points), result in too many missed cases to exceed the impact of NAT-based strategies in more than 99% of simulations. For settings in which NAT is used to confirm Ag-RDT-negative results, Fig. 2B illustrates the relationship of each model parameter to the probability of a “favourable” simulation. In particular, the availability of NAT, sensitivity of clinical judgment amongst those unable to access NAT, and proportion of cases tested during the acute phase were highly influential. Fig. 2C shows the most influential parameters (NAT availability and clinical judgment) in greater detail, with points in blue showing where an Ag-RDT is favourable. Broadly, the figure illustrates that an Ag-RDT would be favourable in settings of low NAT availability and low sensitivity of clinical judgement: in indicative terms, as long as sensitivity of clinical judgement is <90%, and NAT is available to <85% of patients, the probability of Ag-RDT use being favourable is over 98%.

**Figure 2.**
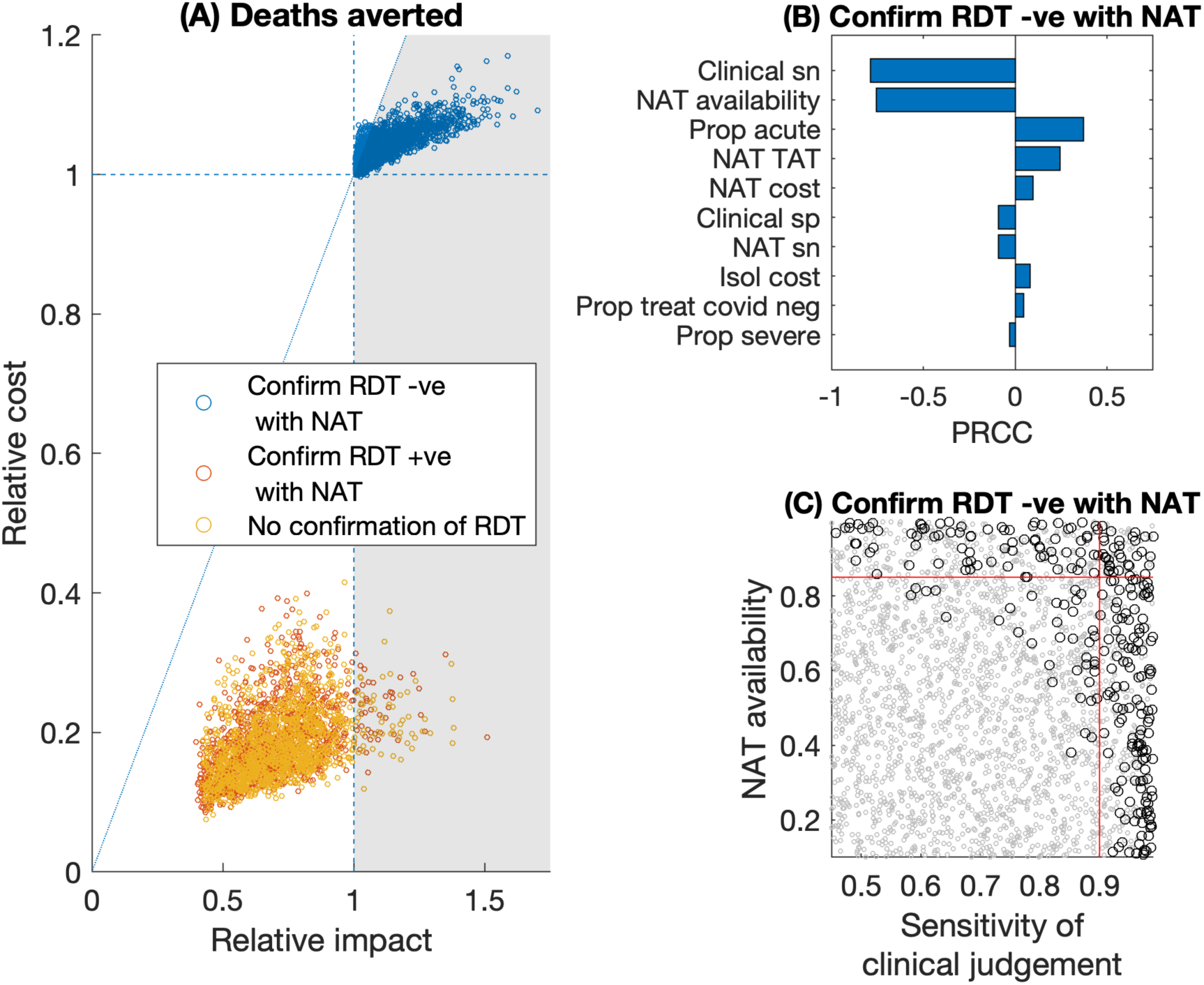
Relative value of Ag-RDT vs NAT testing, for averting deaths in a hospital setting. **(A)** scatter plots for the relative impact of Ag-RDT vs NAT (horizontal axis), vs the relative cost of the two strategies (vertical axis). Each dot represents a single simulation with parameter values drawn from the ranges in table 2. The grey-shaded area shows the region where an Ag-RDT-led strategy would be ‘favourable’ over a NAT-only strategy, meaning that it averts more deaths, and at a lower cost per death averted (see also Fig.S1). Colours of points indicate the adjunctive, confirmatory role of NAT in an Ag-RDT-led strategy (see in-figure legend). Of the blue points, 89% fall in the favourable region. (B) Sensitivity analysis on the blue points in panel (A), to assess when these points fall above, or below, the diagonal dotted reference line. PRCC denotes ‘partial rank correlation coefficient’, against the cost per death averted. (C) The joint role of the two most influential parameters in panel (B). Grey and black points show parameter combinations where an Ag-RDT is favourable, and non-favourable, respectively, relative to NAT. Red lines show 90% sensitivity of clinical judgment (vertical line), and 85% NAT availability (horizontal line). In the lower left quadrant of these lines, an Ag-RDT is favourable over NAT in 93% of simulations. In these results it is assumed that patients are placed in isolation and treated (where indicated) while awaiting a NAT result: Figure S2 in the appendix shows results in the alternative scenario where they are not isolated, nor treated, pending NAT results.

Figure 3 shows similar results in the case of infectious person-days isolated in a hospital setting. Importantly, Fig.3A illustrates the potential for the sole use of Ag-RDT (without NAT confirmation) to offer higher impact at lower cost, than a NAT-based scenario (yellow points, 25% of which lie in the favourable region). Fig. 3C shows a bivariate sensitivity analysis of the two most influential model parameters, demonstrating that an Ag-RDT-only strategy is likely to be favourable in terms of averting infection as long as the sensitivity of clinical judgement in the absence of NAT is <80% and the availability of NAT is <65%. Under these conditions, the probability of Ag-RDT being favourable is over 60%.

**Figure 3.**
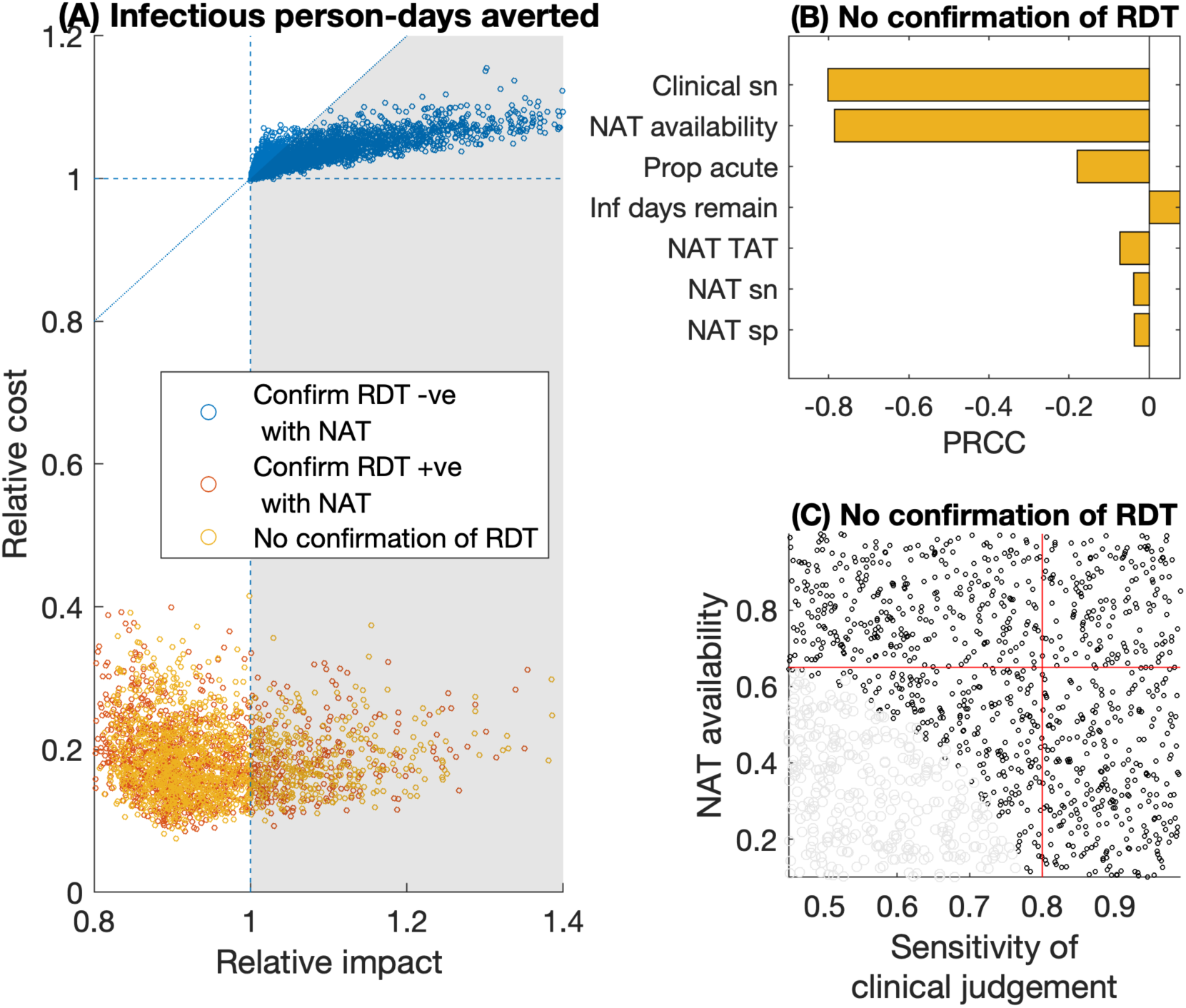
Relative value of Ag-RDT vs NAT testing, for averting infections in a hospital setting. **(A)** scatter plots for the relative impact of Ag-RDT vs NAT (horizontal axis), vs the relative cost of the two strategies (vertical axis). Of the yellow points (no NAT confirmation of Ag-RDT results), 25% fall in the favourable region shaded in grey. Details as in Fig.2A, and Fig.S1. (B) Sensitivity analysis for model parameters on the yellow points in panel (A), to assess when these points fall to the left, or right, of the vertical, dashed reference line. PRCC denotes ‘partial rank correlation coefficient’, against the relative impact of Ag-RDT vs NAT-based strategies. Panel (C) concentrates on the two most influential parameters in this case, NAT availability and sensitivity of clinical judgement. As in Fig.2C, grey and black points show parameter regimes where an Ag-RDT is, respectively, favourable and unfavourable, relative to NAT. Red lines show 75% sensitivity of clinical judgment (horizontal line), and 65% NAT availability (vertical line). In the lower left quadrant of these lines, an Ag-RDT is favourable over NAT in 68% of simulations. In these results it is assumed that patients are placed in isolation while awaiting a NAT result: Figure S3 in the appendix shows results in the alternative scenario where they are not isolated.

These results assume that all patients are placed in isolation while awaiting NAT results, and that those thought to need treatment are initiated on treatment during this time. Figs. S2 and S3 show corresponding results when assuming no isolation nor interim treatment pending NAT results, illustrating that the results are essentially unchanged for deaths averted (Fig. S2). For infectious patient-days isolated, a practice of isolating patients pending NAT results tends to mitigate the drawbacks of multi-day NAT turnaround times: a decision not to isolate pending results therefore reduces the impact of a NAT-based strategy, thus making an Ag-RDT-only strategy more favourable in comparison. Fig. S3 illustrates that, in such a scenario, 89% of simulations place the Ag-RDT strategy in the favourable region.

### Community Screening

Finally, Fig.4 shows results for the community screening scenario. Key assumptions, compared to the hospital scenario, include: the sole priority is to avert infection, because mortality risk in the individuals being evaluated is low; lower prevalence of SARS-CoV-2 amongst those being tested (5%); and we assume individuals are not placed in isolation while awaiting NAT results, because of the infeasibility of doing so. Similarly to Fig.3, confirming Ag-RDT-negative cases with NAT is highly likely to avert more potential transmission than NAT alone, and at lower cost per infectious day averted (blue points, favourable in 100% of simulations). It is also possible for the sole use of Ag-RDT to be more impactful than NAT while costing less (yellow points, favourable in 99% of simulations). Fig. 4B illustrates that key drivers that would tend to increase the relative impact of Ag-RDT-only vs NAT-based strategies are: a higher proportion of individuals that are still in their acute (infectious) phase while being tested; a lower availability of NAT; and a lower sensitivity of clinical judgement. Because the vast majority of simulations place the Ag-RDT in the favourable region, we do not present the bivariate sensitivity analyses that was necessary for the hospital setting scenarios (Figs. 2C, 3C).

**Figure 4.**
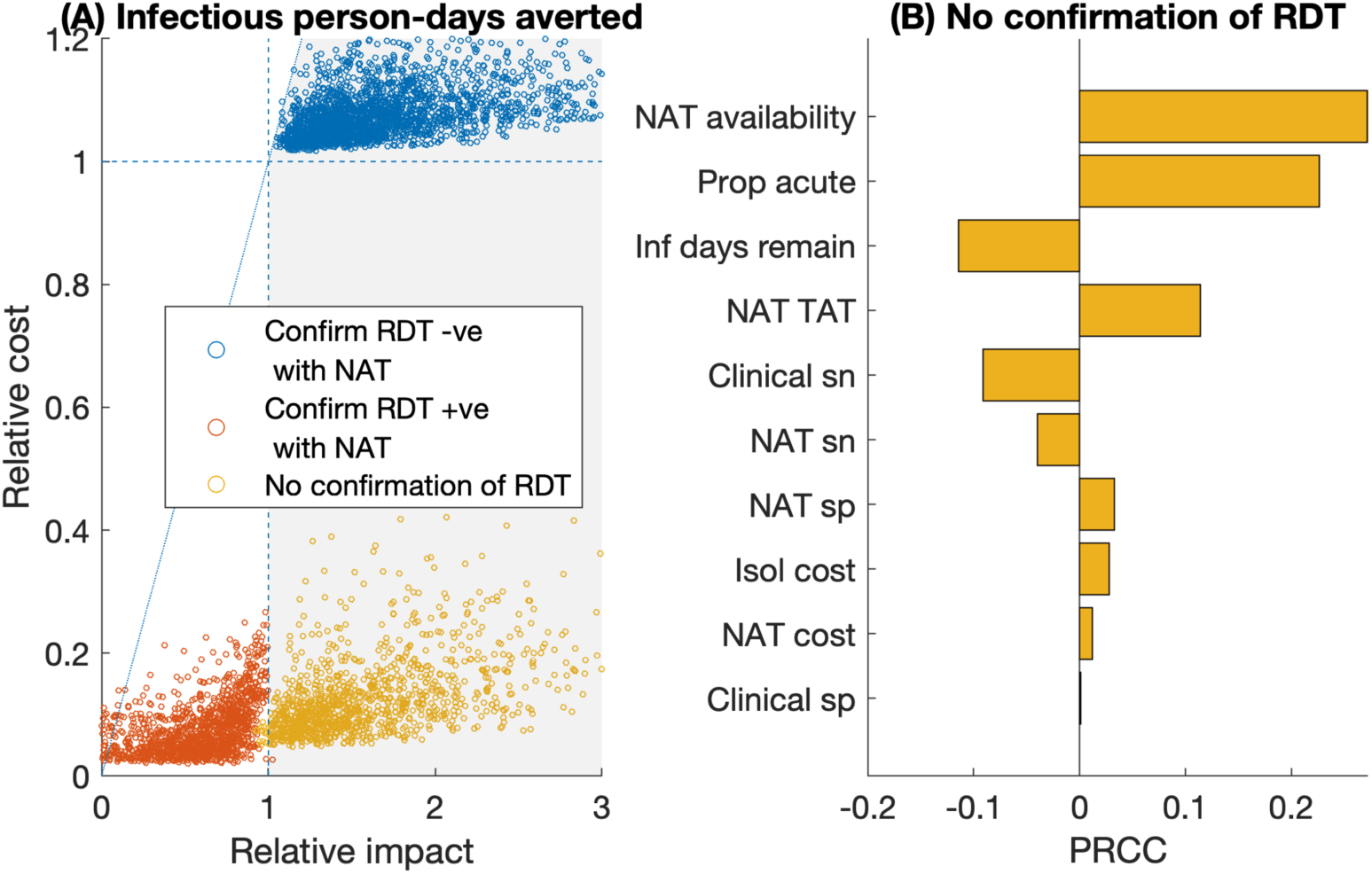
Relative value of Ag-RDT vs NAT testing in a community setting. We assume that in a community setting, the focus is on averting infection, and that any severe cases of respiratory disease are more likely to present in hospital settings (Fig. 3). Hence in this setting, we focus on infectious person-days averted; we also assume that individuals awaiting NAT results are not isolated during this time, owing to the infeasibility of doing so in this setting. (A) Scatter plot of the relative impact of Ag-RDT vs NAT (horizontal axis) vs the relative cost of Ag-RDT vs NAT (vertical axis). Dashed reference lines are as explained in Fig. 2, and in Fig. S1. Of the yellow points (no NAT confirmation of Ag-RDT results), 98% fall in the favourable region shaded in grey. (B) Subgroup sensitivity analysis of the yellow points in panel (A), to assess key factors determining the relative impact of Ag-RDT vs NAT-based strategies. Because the vast majority (98%) of simulations show Ag-RDT being favourable to NAT in this scenario, we do not conduct additional bivariate sensitivity analyses as for Figs. 2C, 3C.

## Discussion

The emergence of antigen-detection rapid diagnostic tests for SARS-CoV-2 has raised important questions about trade-offs between accessibility and performance. Recent models and commentaries have highlighted the potential utility of high-frequency, low-sensitivity testing of asymptomatic individuals (22), and the current analysis demonstrates that under certain circumstances, a less-sensitive but more-accessible test may be preferable for diagnosis of symptomatic COVID-19 as well. In this work we have sought to quantify these trade-offs in a systematic way, using a simple model-based approach to cast light on the situations in which an Ag-RDT, of given performance, may be favoured over NAT. Rather than aiming to specify parameter values with precision, our approach instead embraces parameter uncertainty, by modelling a broad range of scenarios or contextual factors. This approach partly reflects the uncertainty in model parameters, but also their anticipated variability across different country settings and as local epidemics change over time. By structuring our approach in this fashion, we sought to identify the contextual factors that are most important in deciding the value of an Ag-RDT.

Our results suggest that the value of an Ag-RDT-led strategy is strongly supported for evaluating symptomatic individuals in community settings, being highly likely to be simultaneously less costly and more impactful than relying on NAT and clinical judgement (Fig. 4). In hospital settings, the favourability of Ag-RDT may be subject to certain qualifications. For example, in averting deaths, an Ag-RDT, supported by NAT to confirm Ag-RDT-negative results, is likely to be favourable (averting more deaths, at less cost per death averted) to NAT and clinical judgment alone, in settings where NAT is available for less than 85% of patients, and sensitivity of clinical judgement (in the absence of NAT) is less than 90% (Fig. 2).

We note that in the community setting in particular, any reliance on NAT-based testing would face substantial challenges in practice. For example, in most settings it is unlikely that individuals would be adequately isolated while awaiting NAT results, given the large number of unnecessary isolations, and associated burden on patients and families, that such a strategy would incur. Moreover, it will also typically be infeasible to offer timely NAT to all individuals with potential COVID-19 symptoms, given the attendant financial, human resource and supply constraints. In this setting, our analysis shows how an affordable, rapid test, even one with lower performance than NAT, can achieve greater impact overall, and at lower cost, than a strategy that relies on NAT instead.

Notably in both hospital and community scenarios, the key determining factors for the value of an Ag-RDT (namely, the availability of NAT, sensitivity of clinical judgment, and proportion of cases tested during the acute phase) all relate to the ability of the existing system to detect cases of SARS-CoV-2. These findings highlight the potential value of implementation studies to gather data on these factors when making programmatic decisions for the introduction and implementation of new Ag-RDTs in any given setting. Overall, this work serves broadly to illustrate an analytical framework that could be readily adjusted to local realities in different settings. A simple, user-friendly web-based tool is available, to perform the simulations shown here, but also to allow these simulations to be extended to alternative, user-specified parameter ranges (https://covid-ag-rdt.shinyapps.io/model/).

Certain limitations of scope bear mention. Our focus in this work is on identifying the circumstances in which an Ag-RDT might be most valuable, given a pre-specified performance profile. Recent guidance published by WHO addresses target product profiles for Ag-RDTs: that is, how a test should best be optimised in terms of accuracy, cost and ease-of-use, for specified use cases (11). For simplicity, our approach treats transmission-related impact of testing as being directly proportional to the number of days for which testing results in isolation of an infectious person, without considering variation between individuals or over time in the degree of infectivity or the strictness of isolation. Similarly, our assessment of mortality outcomes does not account for the potential of a test to indirectly reduce incidence and mortality by interrupting transmission. Further work using dynamic models of SARS-CoV-2 transmission would be valuable in addressing this gap. In addition, while our results are based on a broad sensitivity analysis, it should be noted that these same results may depend on the range of parameters that we have assumed, and indeed these ranges may vary across different settings. Our user-friendly tool allows users to adapt some of these ranges to specific settings. Amongst other limitations, we have adopted several simplifications, perhaps most importantly assuming a dichotomy between ‘acute’ and ‘recent’ infection and the detectability of each by NAT or Ag-RDT. This assumption ignores potentially important complexities, including how infectivity varies over the clinical course; the stage in the clinical course at which individuals are likely to be tested; and the implications of changing viral/antigen/RNA load over the clinical course, for the ability of a given test to detect infection (16,23–26). Previous modelling studies have incorporated some combinations of these factors (3,22), but longitudinal data on all of these factors will be critical in refining these and other modelling approaches, to account fully for their potential interactions.

In summary, given the immediate importance of virological testing for the control of SARS-CoV-2, it is important for decisions about testing strategy to be guided by the available evidence. Our results show how, in certain clinical conditions, the use of Ag-RDTs could achieve equal or greater impact, and at lower cost, than relying on NAT alone. While the accuracy of diagnostic tools is important, other considerations are also critical: as control efforts increasingly shift from blanket lockdowns towards intensive testing and early identification, the speed, affordability and ease-of-use of diagnostic tools are likely to play an increasingly key role in the response to SARS-CoV-2. Our findings illustrate where such rapid and affordable tests are likely to improve outcomes, at more favourable cost-effectiveness ratios, than reliance on NAT and clinical judgment alone.

## Data Availability

All data used in this study is available in the manuscript and supporting information.

