## Supplementary Information for "Quantifying the potential value of antigen-detection rapid diagnostic tests for COVID-19: a modelling analysis"

#### 1. Model equations

**Number of positive diagnoses**

***NAT-based strategy***

$\boldsymbol{D_{p\_NAT}} =$

| | |
|---|---|
| $[P \times p_{NAT} \times N_{sn}] +$ | *1, True positive (NAT)* |
| $[P \times (1 - p_{NAT}) \times C_{sn}] +$ | *2, True positive (clinical judgement)* |
| $[(1 - P) \times p_{NAT} \times (1 - N_{sp})] +$ | *3, False positive (NAT)* |
| $[(1 - P) \times (1 - p_{NAT}) \times (1 - C_{sp})]$ | *4, False positive (clinical judgement)* |

***Ag-RDT-led strategy***

*Framework:*

$\boldsymbol{D_{p\_RDT}} = [D_{p\_RDT\_only} \times (1 - I_{neg} - I_{pos})] + [D_{p\_RDT\_neg} \times I_{neg}] + [D_{p\_RDT\_pos} \times I_{pos}]$

- *Ag-RDT only:* $(D_{p\_RDT\_only})$

$\boldsymbol{D_{p\_RDT\_only}} =$

| | |
|---|---|
| $[P \times p_{inf} \times R_{\mathrm{inf}\ \_sn}] +$ | *1, True positive, infectious* |
| $[P \times (1 - p_{inf}) \times R_{\mathrm{non\_inf}\ \_sn}] +$ | *2, True positive, non- infectious* |
| $[(1 - P) \times (1 - R_{sp})]$ | *3, False positive* |

- *Confirm Ag-RDT negative with NAT:* $(D_{p\_RDT\_neg})$

$\boldsymbol{D_{p\_RDT\_neg}} =$

| | |
|---|---|
| $[P \times p_{inf} \times R_{\mathrm{inf}\ \_sn}] +$ | *1, True positive (Ag-RDT), infectious* |
| $[P \times (1 - p_{\mathrm{inf}}) \times R_{\mathrm{non\_inf}\ \_sn}] +$ | *2, True positive (Ag-RDT), non-infectious* |
| $[P \times p_{inf} \times (1 - R_{\mathrm{inf}\ \_sn}) \times\ p_{NAT} \times N_{sn}] +$ | *3, True positive (NAT), infectious* |
| $[P \times (1 - p_{inf}) \times (1 - R_{\mathrm{non\_inf}\ \_sn}) \times\ p_{NAT} \times N_{sn}] +$ | *4, True positive (NAT), non-infectious* |
| $[P \times p_{inf} \times (1 - R_{\mathrm{inf}\ \_sn}) \times\ (1 - p_{NAT}) \times C_{sn}] +$ | *5, True positive (clinical judgement), infectious* |
| $[P \times (1 - p_{inf}) \times (1 - R_{\mathrm{non\_inf}\ \_sn}) \times\ (1 - p_{NAT}) \times C_{sn}] +$ | *6, True positive (C.J.), non-infectious* |
| $[(1 - P) \times (1 - R_{\mathrm{sp}})] +$ | *7, False positive (Ag-RDT)* |
| $[(1 - P) \times R_{\mathrm{sp}} \times\ p_{NAT} \times (1 - N_{sp})] +$ | *8, False positive (NAT)* |
| $[(1 - P) \times R_{\mathrm{sp}} \times\ (1 - p_{NAT}) \times (1 - C_{sp})]$ | *9, False positive (C.J.)* |

- *Confirm Ag-RDT positive with NAT:* $(D_{p\_RDT\_pos})$

$\boldsymbol{D_{p\_RDT\_pos}} =$

$[P \times p_{inf} \times R_{\text{inf}\_sn} \times N_{sn}] +$ *1, True positive, infectious*
$[P \times (1 - p_{inf}) \times R_{\text{non\_inf}\_sn} \times N_{sn}] +$ *2, True positive, non-infectious*
$[(1 - P) \times (1 - R_{\text{sp}}) \times (1 - N_{sp})]$ *3, False positive*

**<u>Number of false-negative diagnoses</u>**

***NAT-based strategy***

$\boldsymbol{D_{n\_NAT}} =$

$[P \times p_{NAT} \times (1 - N_{sn})] +$ *1, False negative (NAT)*
$[P \times (1 - p_{NAT}) \times (1 - C_{sn})]$ *2, False negative (clinical judgement)*

***Ag-RDT-led strategy***

*Framework*:
$\boldsymbol{D_{n\_RDT}} = [D_{n\_RDT\_only} \times (1 - I_{neg} - I_{pos})] + [D_{n\_RDT\_neg} \times I_{neg}] + [D_{n\_RDT\_pos} \times I_{pos}]$

- *Ag-RDT only:* $(D_{n\_RDT\_only})$

$\boldsymbol{D_{n\_RDT\_only}} =$

$[P \times p_{inf} \times (1 - R_{\text{inf}\_sn})] +$ *1, False negative, infectious*
$[P \times (1 - p_{inf}) \times (1 - R_{\text{non\_inf}\_sn})]$ *2, False negative, non-infectious*

- *Confirm Ag-RDT negative with NAT:* $(D_{n\_RDT\_neg})$

$\boldsymbol{D_{n\_RDT\_neg}} =$

$[P \times p_{inf} \times (1 - R_{\text{inf}\_sn}) \times p_{NAT} \times (1 - N_{sn})] +$ *1, False negative (Ag-RDT & NAT), infectious*
$[P \times (1 - p_{inf}) \times (1 - R_{\text{non\_inf}\_sn}) \times p_{NAT} \times (1 - N_{sn})] +$ *2, False neg. (Ag-RDT & NAT), non-inf*
$[P \times p_{inf} \times (1 - R_{\text{inf}\_sn}) \times (1 - p_{NAT}) \times (1 - C_{sn})] +$ *3, False neg. (Ag-RDT & C.J.), inf*
$[P \times (1 - p_{inf}) \times (1 - R_{\text{non\_inf}\_sn}) \times (1 - p_{NAT}) \times (1 - C_{sn})]$ *4, False neg. (Ag-RDT & C.J), non-inf*

- *Confirm Ag-RDT positive with NAT:* $(D_{n\_RDT\_pos})$

$\boldsymbol{D_{n\_RDT\_pos}} =$

$[P \times p_{inf} \times (1 - R_{\text{inf}\_sn})] +$ *1, False negative (missed by Ag-RDT), infectious*
$[P \times (1 - p_{inf}) \times (1 - R_{\text{non\_inf}\_sn})] +$ *2, False negative (Ag-RDT), non- infectious*
$[P \times p_{inf} \times R_{\text{inf}\_sn} \times (1 - N_{sn})] +$ *3, False negative (NAT), infectious*
$[P \times (1 - p_{inf}) \times R_{\text{non\_inf}\_sn} \times (1 - N_{sn})]$ *4, False negative (NAT), non-infectious*

### Number of NAT tests needed

#### *NAT-based strategy*

$\boldsymbol{N_{NAT}} = p_{NAT}$

#### *Ag-RDT-led strategy*

*Framework*:
$\boldsymbol{N_{RDT}} = [N_{RDT\_only} \times (1 - I_{neg} - I_{pos})] + [N_{RDT\_neg} \times I_{neg}] + [N_{RDT\_pos} \times I_{pos}]$

- *Ag-RDT only:* $(N_{RDT\_only})$

$\boldsymbol{N_{RDT\_only}} = 0$

- *Confirm Ag-RDT negative with NAT:* $(N_{RDT\_neg})$

$\boldsymbol{N_{RDT\_neg}} =$

$[P \times p_{inf} \times (1 - R_{\mathrm{inf}\ \_sn}) \times\ p_{NAT}] +$ *1, False negative (missed by Ag-RDT), infectious*
$[P \times (1 - p_{inf}) \times (1 - R_{\mathrm{non\_inf}\ \_sn}) \times\ p_{NAT}] +$ *2, False negative (Ag-RDT), non- infectious*
$[(1 - P) \times R_{\mathrm{sp}} \times\ p_{NAT}]$ *3, True negative (Ag-RDT)*

- *Confirm Ag-RDT positive with NAT:* $(N_{RDT\_pos})$

$\boldsymbol{N_{RDT\_pos}} =$

$[P \times p_{inf} \times R_{\mathrm{inf}\ \_sn}] +$ *1, True positive (detected by Ag-RDT), infectious*
$[P \times (1 - p_{inf}) \times R_{\mathrm{non\_inf}\ \_sn}] +$ *2, True positive (Ag-RDT), non-infectious*
$[(1 - P) \times (1 - R_{\mathrm{sp}})]$ *3, False positive (Ag-RDT)*

### Number of Ag-RDT tests needed

#### *NAT-based strategy*

$\boldsymbol{R_{NAT}} = 0$

#### *Ag-RDT-led strategy*

*Framework*:
$\boldsymbol{R_{RDT}} = [R_{RDT\_only} \times (1 - I_{neg} - I_{pos})] + [R_{RDT\_neg} \times I_{neg}] + [R_{RDT\_pos} \times I_{pos}]$

- *Ag-RDT only:* $(R_{RDT\_only})$

$\boldsymbol{R_{RDT\_only}} = 1$

- *Confirm Ag-RDT negative with NAT:* $(R_{RDT\_neg})$

$\boldsymbol{R_{RDT\_neg}} = 1$

- *Confirm Ag-RDT positive with NAT:* $(R_{RDT\_pos})$

$\boldsymbol{R_{RDT\_pos}} = 1$

**Number of deaths per person**

*Framework for all algorithms:*

$$\boldsymbol{deaths} =$$
$$\left[True\ positives\ \times p_{severe} \times p_{treat_{pos}} \times M_{treat}\right] +$$
$$\left[True\ positives\ \ \times p_{severe} \times \left(1 - p_{treat_{pos}}\right) \times M_{no_{treat}}\right] +$$
$$[False\ negatives\ \times p_{severe} \times M_{no_{treat}}]$$

**Number of infectious days per person**

*Framework for all algorithms, except Ag-RDT only:*

$$\boldsymbol{ID} =$$
$$[True\ positives\ diagnosed\ by\ NAT\ \times p_{inf} \times \min(\ D_{inf}, D_{NAT}) \times (1 - N_{isol})] +$$
$$\left[False\ negatives, NAT\ \times p_{inf} \times \left(\left(\min(\ D_{inf}, D_{NAT}) \times (1 - N_{isol})\right) + \max\left(D_{inf} - D_{NAT}, 0\right)\right)\right] +$$
$$[False\ negatives, clinical\ judgement \times p_{inf} \times D_{inf}]$$

*Framework for Ag-RDT only:*

$$\boldsymbol{ID} = [D_{n\_RDT\_only} \times D_{inf}]$$

**Cost per person**

***Framework for all algorithms:***

$$Cost_{total} = Cost_{testing} + Cost_{isolation} + Cost_{treatment}$$

***NAT-based strategy***

- *Cost of testing*

$$Cost_{NAT\_testing} = N_{NAT} \times C_{NAT}$$

- *Cost of isolation*

$$Cost_{NAT\_isolation} =$$

$$[p_{NAT} \times N_{isol} \times D_{NAT} \times C_{isol}] +$$
$$\left[\left(D_{p\_NAT}(1) + D_{p\_NAT}(3)\right) \times N_{isol} \times \max(D_{isol} - D_{NAT}, 0) \times C_{isol}\right] +$$
$$[(D_{p\_NAT}(1) + D_{p\_NAT}(3)) \times (1 - N_{isol}) \times D_{isol} \times C_{isol}] +$$
$$[(D_{p\_NAT}(2) + D_{p\_NAT}(4)) \times D_{isol} \times C_{isol}]$$

- *Cost of treatment*

$$Cost_{NAT\_treatment} =$$

$$\left[P \times p_{NAT} \times N_{isol} \times p_{severe} \times p_{treat_{pos}} \times D_{NAT} \times C_{treat}\right] +$$
$$\left[(1 - P) \times p_{NAT} \times N_{isol} \times p_{treat_{neg}} \times D_{NAT} \times C_{treat}\right] +$$
$$[D_{p\_NAT}(3) \times (1 - N_{isol}) \times p_{treat_{neg}} \times D_{treat} \times C_{treat}] +$$
$$\left[D_{p\_NAT}(3) \times N_{isol} \times p_{treat_{neg}} \times \max(D_{treat} - D_{NAT}, 0) \times C_{treat}\right] +$$
$$\left[D_{p\_NAT}(4) \times p_{treat_{neg}} \times D_{treat} \times C_{treat}\right] +$$

$[D_{p\_NAT}(1) \times (1 - N_{isol}) \times p_{severe} \times p_{treat_{pos}} \times D_{treat} \times C_{treat}] +$
$\left[D_{p\_NAT}(1) \times N_{isol} \times p_{severe} \times p_{treat_{pos}} \times \max(D_{treat} - D_{NAT}, 0) \times C_{treat}\right] +$
$[D_{p\_NAT}(2) \times p_{severe} \times p_{treat_{pos}} \times D_{treat} \times C_{treat}]$

***Ag-RDT only***

- *Cost of testing*

$Cost_{RDT\_testing} = C_{RDT}$

- *Cost of isolation*

$Cost_{RDT\_isolation} = D_{p\_RDT\_only} \times D_{isol} \times C_{isol}$

- *Cost of treatment*

$Cost_{RDT\_treatment} =$

$[(D_{p\_RDT\_only}(1) + D_{p\_RDT\_only}(2)) \times p_{severe} \times p_{treat_{pos}} \times D_{treat} \times C_{treat}] +$
$[FD_{p\_RDT\_only}(3) \times p_{treat_{neg}} \times D_{treat} \times C_{treat}]$

***Confirm Ag-RDT negative with NAT***

- *Cost of testing*

$Cost_{RDT\_neg\_testing} = C_{RDT} + [N_{RDT\_neg} \times C_{NAT}]$

- *Cost of isolation*

$Cost_{RDT\_neg\_isolation} =$

$[(D_{p\_RDT\_neg}(1) + D_{p\_RDT\_neg}(2) + D_{p\_RDT\_neg}(7) + D_{p\_RDT\_neg}(5) + D_{p\_RDT\_neg}(6)$
$+ D_{p\_RDT\_neg}(9)) \times D_{isol} \times C_{isol}] +$
$[N_{RDT\_neg} \times N_{isol} \times D_{NAT} \times C_{isol}] +$
$[(D_{p\_RDT\_neg}(3) + D_{p\_RDT\_neg}(4) + D_{p\_RDT\_neg}(8)) \times N_{isol} \times \max(D_{isol} - D_{NAT}, 0) \times C_{isol}] +$
$[(D_{p\_RDT\_neg}(3) + D_{p\_RDT\_neg}(4) + D_{p\_RDT\_neg}(8)) \times (1 - N_{isol}) \times D_{isol} \times C_{isol}]$

- *Cost of treatment*

$Cost_{RDT\_neg\_treatment} =$

$\left[(N_{RDT\_neg}(1) + N_{RDT\_neg}(2)) + \times N_{isol} \times p_{severe} \times p_{treat_{pos}} \times D_{NAT} \times C_{treat}\right] +$
$\left[(D_{p\_RDT\_neg}(1) + D_{p\_RDT\_neg}(2) + D_{p\_RDT\_neg}(5) + D_{p\_RDT\_neg}(6)) \times p_{severe} \times p_{treat_{pos}} \times D_{treat}\right.$
$\left.\times C_{treat}\right] +$
$\left[(D_{p\_RDT\_neg}(3) + D_{p\_RDT\_neg}(4)) \times N_{isol} \times p_{severe} \times p_{treat_{pos}} \times \max(D_{treat} - D_{NAT}, 0) \times C_{treat}\right]$
$+$
$[(D_{p\_RDT\_neg}(3) + D_{p\_RDT\_neg}(4)) \times (1 - N_{isol}) \times p_{severe} \times p_{treat_{pos}} \times D_{treat} \times C_{treat}] +$
$\left[N_{RDT\_neg}(3) \times N_{isol} \times p_{treat_{neg}} \times D_{NAT} \times C_{treat}\right] +$
$\left[(D_{p\_RDT\_neg}(7) + D_{p\_RDT\_neg}(9)) \times p_{treat_{neg}} \times D_{treat} \times C_{treat}\right] +$
$\left[D_{p\_RDT\_neg}(8) \times N_{isol} \times p_{treat_{neg}} \times \max(D_{treat} - D_{NAT}, 0) \times C_{treat}\right] +$
$[D_{p\_RDT\_neg}(8) \times (1 - N_{isol}) \times p_{treat_{neg}} \times D_{treat} \times C_{treat}]$

***Confirm Ag-RDT positive with NAT***

- *Cost of testing*

$Cost_{RDT\_pos\_testing} = C_{RDT} + [N_{RDT\_pos} \times C_{NAT}]$

- *Cost of isolation*

$Cost_{RDT\_pos\_isolation} =$

$\left[N_{RDT\_pos} \times N_{isol} \times D_{NAT} \times C_{isol}\right] +$
$\left[D_{p\_RDT\_pos} \times N_{isol} \times \max(D_{isol} - D_{NAT}, 0) \times C_{isol}\right] +$
$[D_{p\_RDT\_pos} \times (1 - N_{isol}) \times D_{isol} \times C_{isol}]$

- *Cost of treatment*

$Cost_{RDT\_pos\_treatment} =$

$\left[(N_{RDT\_pos}(1) + N_{RDT\_pos}(2)) \times N_{isol} \times p_{severe} \times p_{treat_{pos}} \times D_{NAT} \times C_{treat}\right] +$
$\left[\left(D_{p\_RDT\_pos}(1) + D_{p\_RDT\_pos}(2)\right) \times N_{isol} \times p_{severe} \times p_{treat_{pos}} \times \max(D_{treat} - D_{NAT}, 0) \times C_{treat}\right]$
$+$
$[\left(D_{p\_RDT\_pos}(1) + D_{p\_RDT\_pos}(2)\right) \times (1 - N_{isol}) \times p_{severe} \times p_{treat_{pos}} \times D_{treat} \times C_{treat}] +$
$\left[N_{RDT\_pos}(3) \times N_{isol} \times p_{treat_{neg}} \times D_{NAT} \times C_{treat}\right] +$
$\left[D_{p\_RDT\_pos}(3) \times N_{isol} \times p_{treat_{neg}} \times \max(D_{treat} - D_{NAT}, 0) \times C_{treat}\right] +$
$[D_{p\_RDT\_pos}(3) \times (1 - N_{isol}) \times p_{treat_{neg}} \times D_{treat} \times C_{treat}]$

### 2. Expert consultation

To identify appropriate use cases for implementation of antigen-detection RDTs for SARS-CoV-2, we conducted a series of four expert consultations. For each consultation, we identified an in-country expert with knowledge of the landscape of diagnostic testing for infectious diseases and the country's response to the COVID-19 pandemic. We consulted experts in Brazil, India, Nigeria, and South Africa (experts listed in Table S1, below) – countries with large populations, a breadth of income levels, broad geographic representation, and COVID-19 epidemics of substantial magnitude. Consultations were performed between July 2 and 7, 2020. Each consultation was structured as follows:

First, we provided background on the potential characteristics of an Ag-RDT: results within 30 minutes, more feasible to perform at point of care, performed on oropharyngeal/nasal/salivary specimens, and less sensitive than nucleic acid amplification tests. Second, we described the primary research objective: to estimate the benefits and harms of implementing Ag-RDTs in specific use cases. Third, we provided examples of paradigmatic use cases, in terms of eligible population and intended use of the Ag-RDT. Fourth, we asked in-country experts to identify at least two important use cases for their country's setting, asking that they define: (a) the eligible population; (b) the intended use/incremental benefit of Ag-RDT; (c) the status quo – how members of the population would be managed in the absence of an Ag-RDT; (d) how the status quo would change in the presence of an Ag-RDT (for both people testing positive and negative on Ag-RDT); (e) the most important outcomes/measures of value; and (f) the most important tradeoffs to consider.

Each consultation lasted between 60 and 90 minutes, resulting in the following use cases being identified:

- Community-based testing of symptomatics, in decentralised clinics or in dedicated testing facilities in containment zones (India and South Africa)
- Testing of symptomatic individuals in health facilities and amongst those being admitted to hospital (India and South Africa)
- Testing of asymptomatic employees contacting high-risk populations such as long-term care facilities (South Africa)
- Testing of symptomatic outpatients (Brazil and Nigeria)
- Testing of asymptomatic employees in high-risk occupations such as healthcare workers (Brazil and Nigeria)

For the purpose of this analysis, we focused on the first two use cases listed above. However, the approach described in our study can also straightforwardly be extended to the other use cases listed here.

| **Name of expert** | **Country** | **Affiliation** |
| --- | --- | --- |
| Kiran Rade | India | Indian Council of Medical Research |
| Dhamari Naidoo | Nigeria | WHO Country Office |
| Francois Venter | South Africa | WITS Reproductive Health and HIV Institute |
| Amilcar Tanuri | Brazil | Federal University of Rio de Janeiro |

**Table S1. List of country experts consulted to identify appropriate use cases, for an Ag-RDT**

### 3. Incremental cost-effectiveness ratios

| Testing strategy | | Hospital setting | | Community setting |
|---|---|---|---|---|
| | | **Cost per death averted ($)** | **Cost per infectious person-day isolated ($)** | **Cost per infectious person-day isolated ($)** |
| **NAT-based** | | $119,830 (32,080-491,070) | $214 (67-571) | $732 (179-2541) |
| **Ag-RDT-led** | No Ag-RDT confirmation | $43,850 (13,100-172,670) | $58 (18-156) | $88 (26-227) |
| | Confirm Ag-RDT -ve with NAT | $113,130 (29,470-451,600) | $205 (63-531) | $463 (128-1492) |
| | Confirm Ag-RDT +ve with NAT | $45,750 (14,600-173,750) | $59 (20-156) | $185 (45-651) |

**Table S2. Summary of incremental cost-effectiveness ratios of results presented in the main text**. As explained in the Methods, impact and cost are estimated relative to a baseline of no testing, and no intervention. Numbers in brackets give 95% uncertainty intervals.

### 4. Additional supplementary Figures

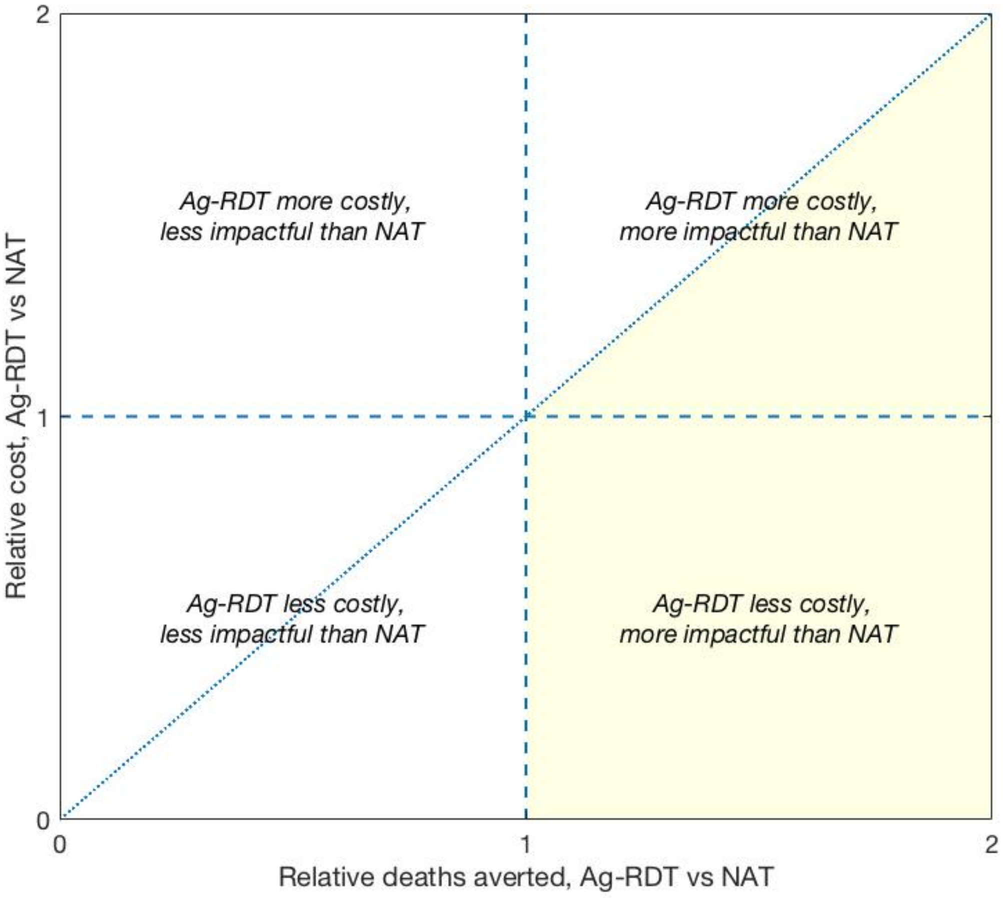


**Figure S1. Schematic illustration for visualising the value of an Ag-RDT-led strategy**, relative to a scenario involving NAT and clinical judgement. Although the figure involves deaths averted, the same structure applies for averting infectious person-days. For a given set of parameters drawn from the parameter ranges shown in Table 2, we simulate the cost and impact of a given Ag-RDT-led strategy, and of a NAT-based testing strategy, both relative to a no-intervention scenario. This outcome is then represented in the figure by plotting the relative deaths averted by Ag-RDT vs NAT (horizontal axis) against the relative cost of the two strategies (vertical axis). Thus, for example, in the lower right quadrant, an Ag-RDT-led strategy would cost less, but have more impact, than NAT. The diagonal dashed line shows an important threshold: for points below this line, an Ag-RDT-led strategy would cost less per death averted than NAT, and vice versa. Overall, therefore, the shaded area shows the region in which an Ag-RDT would simultaneously cost less per death averted, and avert more deaths overall, than NAT. We denote this area as the 'favourable region' for an Ag-RDT, and elsewhere as 'non-favourable': in our current analysis we aim to identify the circumstances under which an Ag-RDT, of a given performance and cost, would occupy this region.

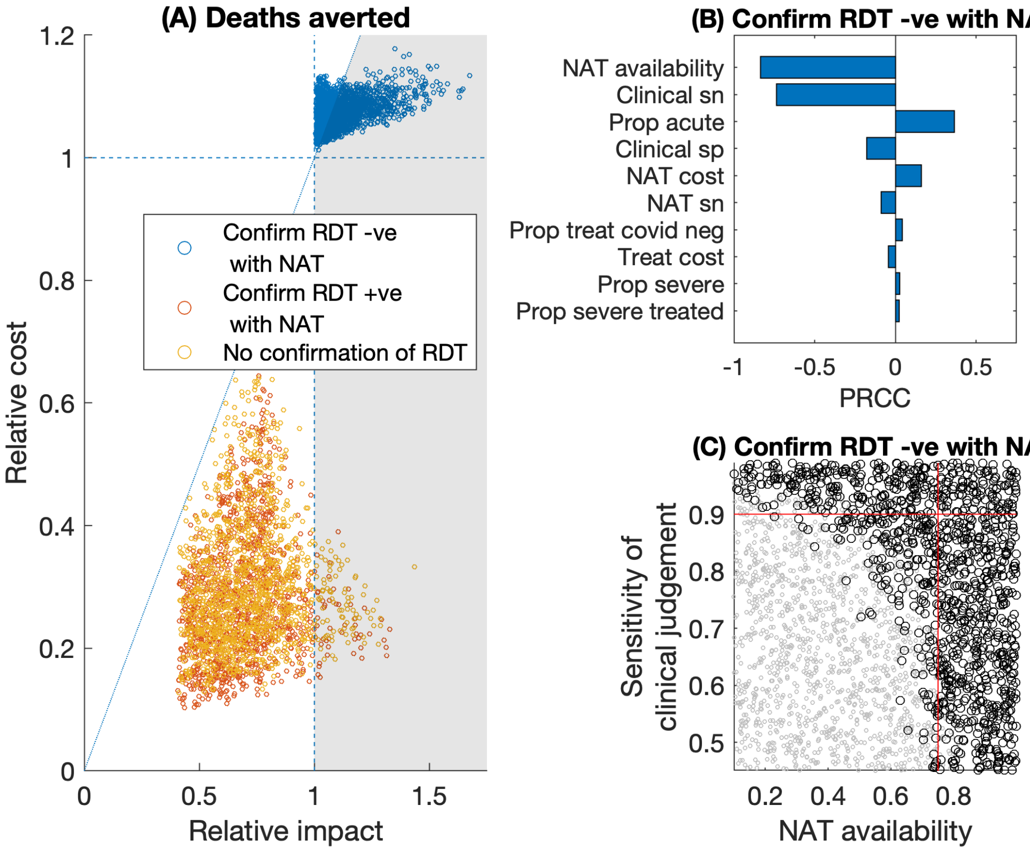


**Figure S2. Relative value of Ag-RDT-led vs NAT-based testing, for averting deaths in a hospital setting.** The figure shows the same results as those presented in Fig.2 in the main text, but here assuming that all patients awaiting a NAT result (whether as part of a NAT-based strategy or for confirmation of Ag-RDT results) are *not* isolated during this time, nor are the more severe cases initiated on treatment. Results illustrate qualitatively similar findings to those shown in the main text. In panel (A), in the scenario where Ag-RDT-negative results are confirmed using NAT (blue points), 55% of simulations place the Ag-RDT-led strategy in the favourable region, below the diagonal dashed line. Panels (B, C) show additional sensitivity analyses for these points in particular, as described in Fig.2. In (C), red lines show 75% NAT availability (vertical line), and 90% sensitivity of clinical judgment (horizontal line). In the lower left quadrant of these lines, an Ag-RDT is favourable over NAT in 86% of simulations.

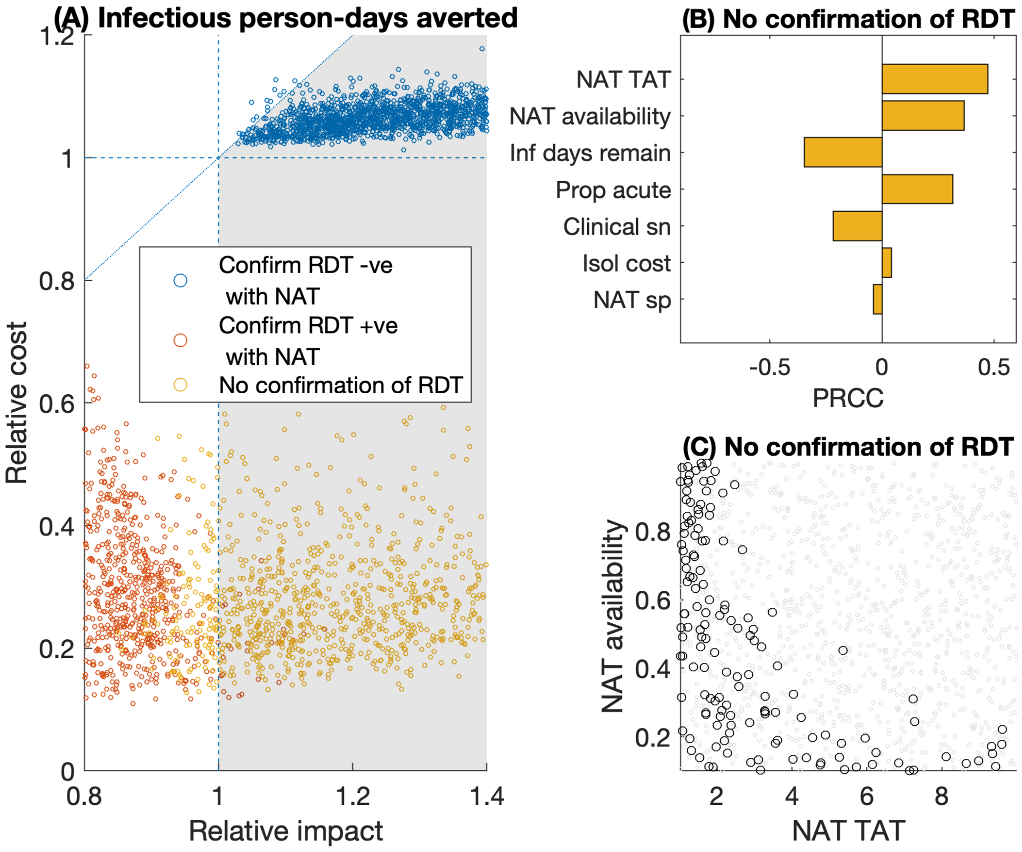


**Figure S3. Relative value of Ag-RDT-led vs NAT-based testing, for averting infections in a hospital setting.** The figure shows the same results as those presented in Fig.3 in the main text, but here assuming that all patients awaiting a NAT result (whether as part of a NAT-based strategy or for confirmation of Ag-RDT results) are not isolated during this time, nor are the more severe cases initiated on treatment. In panel (A), in the scenario where there is no NAT confirmation of Ag-RDT results (yellow points), 89% of simulations place the Ag-RDT-led strategy in the favourable region, to the right of the vertical, dashed line. Panels (B, C) show additional sensitivity analyses for these points in particular, as described in Fig.3. In panel (C), because an Ag-RDT is favourable in the vast majority of simulations, here we do not present illustrative thresholds for the two parameters shown.
